# SARS-CoV-2 Pandemic Non-Pharmacologic Interventions Temporally Associated with Reduced Pediatric Infections Due to *Mycoplasma pneumoniae* and Co-Infecting Respiratory Viruses in Arkansas

**DOI:** 10.1101/2023.08.05.23293566

**Authors:** Bobby L. Boyanton, Rachel A. Frenner, Ashton Ingold, Lilliam Ambroggio, Joshua L. Kennedy

## Abstract

**Introduction:** Non-pharmacologic interventions (NPIs), such as universal masking, implemented during the SARS-CoV-2 pandemic have reduced respiratory infections among children. This study focuses on evaluating the impact of NPIs on *Mycoplasma pneumoniae* infections in children, analyzing data from two hospitals in Arkansas, and examining age-related differences and coinfections with other viruses.

**Methods:** The study was approved by the Institutional Review Board and included patients aged ≤18 years with upper respiratory tract symptoms. Data from the FilmArray® Respiratory Panel (FARP) were collected and divided into pre-NPI and NPI periods for analysis. Total test positivity rate and interval change in the positivity rate were evaluated. Statistical differences were determined by Chi-square (χ^2^-independence) analysis.

**Results:** A total of 68,949 tests were performed with a statistical increase in testing during the NPI period. The overall test positivity rate for *M. pneumoniae* decreased by 74% (0.86% to 0.03%) during the NPI period, and the preschool age group had the highest number of positive tests in the pre- and NPI periods (Pre-NPI: n=40, NPI: n=12 positive tests, p=<0.001). The reduction in *M. pneumoniae* infections was consistent across age groups. Coinfections with other respiratory viruses, particularly human rhinovirus/enterovirus, were observed at much lower levels.

**Conclusions:** NPIs effectively reduced *M. pneumoniae* in pediatric patients in Arkansas, and coinfections with specific viruses still occurred, albeit at lower levels during the SARS-CoV-2 pandemic. As NPIs are relaxed and the pandemic ends, we expect *M. pneumoniae* infections to return to pre-pandemic levels within the next 1-2 years.

## Introduction

Non-pharmacologic interventions (NPIs), such as universal masking, physical distancing, and school closures, were recommended in March 2020 to mitigate the SARS-CoV-2 pandemic.(1) These interventions were implemented to reduce the transmission of the virus and prevent the spread of COVID-19. While the primary focus of NPIs was on preventing COVID-19 cases, additional benefits have been observed with other respiratory infections, especially in children. Compliance with NPIs has resulted in a significant decrease in respiratory infections among children. Multiple studies have now reported reduced rates of respiratory illnesses, such as the common cold, croup, otitis media, pharyngitis, pneumonia, and sinusitis caused by different viruses and bacteria.(2-14) The use of masks has been particularly effective in preventing the spread of respiratory droplets and aerosol particles that can carry infectious agents.

*Mycoplasma pneumoniae* is a bacterium that infects the respiratory epithelium and causes mild to severe upper and lower respiratory tract disease in children and adults, including community-acquired pneumonia (CAP). It is transmitted through person-to-person contact via aerosols produced during coughing and sneezing. Implementing infection prevention measures such as wearing masks and maintaining physical distance can help reduce the transmission of *M. pneumoniae* and other aerosol-spread infectious agents. During the SARS-CoV-2 pandemic, the widespread adoption of NPIs led to a significant decrease in upper and lower respiratory tract infections, including those caused by *M. pneumoniae*. (14-21) This reduction in infections can be attributed to the decreased opportunities for close contact and the adherence to hygiene practices like handwashing and respiratory etiquette.

However, the detection and surveillance of *M. pneumon*iae infections have been challenging due to the lack of standardized diagnostic methods. Diagnostic testing for *M. pneumoniae* includes nucleic acid amplification tests (NAAT), antigen tests, culture, and serology. However, these methods vary in sensitivity and specificity, making it difficult to accurately track the prevalence of these infections. Consequently, surveillance efforts have been limited, and comprehensive data on the impact of NPIs on *M. pneumoniae* infections have been scarce. Furthermore, *M. pneumoniae* infections pose a diagnostic challenge in clinical differentiation of symptoms from other respiratory viral pathogens. Age related differences are also possible with younger patients either showing no noticeable symptoms or presenting with coryza and wheezing, without the presence of fever.

To address this gap in knowledge, a study was conducted at Arkansas Children’s Hospital (ACH) and Arkansas Children’s Northwest (ACNW) to evaluate the impact of NPIs on *M. pneumoniae* infection rates in children. By analyzing the data collected from these two medical facilities, the study aimed to provide valuable insights into the association between NPIs and *M. pneumoniae* infections in children. A secondary goal was to assess age-related differences and coinfections with other viruses in the children of Arkansas before and during the COVID-19 pandemic.

## Methods

This retrospective cross-sectional study received approval from the Institutional Review Board of the University of Arkansas for Medical Sciences with waiver of consent and HIPAA authorization (No. 274080). The study included patients aged ≤18 years who presented with upper respiratory tract symptoms and were tested using the BioFire® FilmArray® Respiratory Panel (FARP; bioMerieux, Durham, NC, USA) from November 1, 2017, to December 31, 2022 at Arkansas Children’s Hospital (Little Rock, AR) or Arkansas Children’s Northwest (Springdale, AR). The FARP detected various respiratory pathogens, including adenovirus, coronaviruses, human metapneumovirus, human rhinovirus/enterovirus, influenza viruses, parainfluenza viruses, respiratory syncytial virus, *Bordetella pertussis, Chlamydia pneumoniae*, and *Mycoplasma pneumoniae*. During the study period, the U.S. Food and Drug Administration approved newer versions of the FARP for clinical use. Specifically, *B. parapertussis* and SARS-CoV-2 were included in December 2017 and June 2020, respectively. Patient demographic information, FARP test results, and data on coinfections were collected from the electronic health record system (EPIC, Verona, WI).

In Arkansas, NPIs were implemented in late March 2020, including universal masking, physical distancing, restricted access to public activities, and temporary closure of in-person school and daycare facilities. Enhanced hand hygiene and surface cleansing were also recommended. NPIs were gradually lifted from March 2021 onwards. Therefore, the data were divided into pre-NPI (November 2017-March 2020) and NPI (April 2020-December 2022) periods for analysis.

Based on previous studies, patients were categorized into age groups: preschool (0-5 years), elementary school (6-10 years), middle school (11-13 years), and high school (14-18 years) (ref). Coinfections with other respiratory pathogens were also evaluated. Descriptive statistics and Chi-square tests were used to calculate the test positivity rate (number of positive tests divided by all FARP performed at ACH) and the change in positivity rate between the pre-NPI and NPI periods. Statistical analysis was performed using Microsoft Excel 365, with significance determined at a P-value of <0.05. Descriptive statistics were used to calculate the test positivity rate (number of positive tests divided by all FARP performed at ACH) and interval change in the positivity rate (pre-NPI minus NPI periods). Statistical differences were determined by Chi-square (χ^2^-independence) analysis. All statistical analyses were performed with Microsoft Excel 365 (Microsoft Corp., Redmond, Washington, USA); *P* values <0.05 were considered statistically significant.

## Results

During the study, a total of 68,949 tests were performed on 42,528 unique patients 18 years old or younger. Among these patients, 46% were female. The tests were divided into two periods: the pre-NPI period (8,972 tests) and the NPI period (59,977 tests). There was a 600% increase in total testing volume during the NPI period when compared to the pre-NPI period. Table 1 provides an overview of the overall and age group-specific test positivity rates and the decrease in test positivity rates between the pre-NPI and NPI periods. In the pre-NPI period, there was a test positivity rate of 0.86% (n=77). In the NPI period, the test positivity rate was 0.03% (n=19). The overall test positivity rate decreased by 74% (p<0.001).

**Table 1.** Summary of Patient Demographics, *M. pneumoniae* Test Positivity Rate, and Interval Change (Percent Change Following NPI Implementation).

| Demographic Information | | Pre-NPI Period (11/2017-03/2020) | | | NPI Period (04/2020-12/2022) | | | Interval Change | $\chi^2$ |
| --- | --- | --- | --- | --- | --- | --- | --- | --- | --- |
| Age (yr.) | Unique Patients | Positive Tests | Total Tests | % Positive | Positive Tests | Total Tests | % Positive | (%) | P value |
| All | 42,528 | 77 | 8,972 | 0.86 | 19 | 59,977 | 0.03 | -74.0 | <.001 |
| 0-5 | 31,597 | 40 | 7,010 | 0.57 | 12 | 45,674 | 0.03 | -70.0 | <.001 |
| 6-10 | 6,311 | 24 | 881 | 2.72 | 5 | 7,623 | 0.07 | -75.0 | <.001 |
| 11-13 | 2,398 | 7 | 441 | 1.59 | 2 | 2,927 | 0.07 | -71.4 | <.001 |
| 14-18 | 3,061 | 6 | 640 | 0.94 | 0 | 3,753 | 0.00 | -100.0 | <.001 |

The preschool age group saw a 70% decrease in *M. pneumoniae* positivity rate, the elementary school group saw a 75% decrease, the middle school group saw a 71.4% decrease, and the high school group saw a 100% decrease (P<0.001) in the NPI period compared to the pre-NPI period. However, the distribution of tests across different age groups remained relatively stable during both periods, with preschoolers accounting for 78±3% and 75±6%, elementary school students accounting for 10±2% and 12±3%, middle school students accounting for 5±1% and 5±1%, and high school students accounting for 7±1% and 7±3% during the pre-NPI and NPI periods, respectively (Figure 1A). The preschool group had the highest number of positive tests for *M. pneumoniae* (n=40 to 12) but the lowest age group-specific percent positivity rate due to the larger number of total tests performed (0.57% to 0.03%). The elementary school group had the second-highest number of positive tests for *M. pneumoniae* (n=24 to 5) and the highest age group-specific percent positivity rate (2.72% to 0.07%). The middle school (n=7 to 2) and high school groups (n=6 to 0) had the fewest number of positive tests for *M. pneumoniae* and the fewest total tests performed (Figure 1B, 1C).

**Figure 1.**
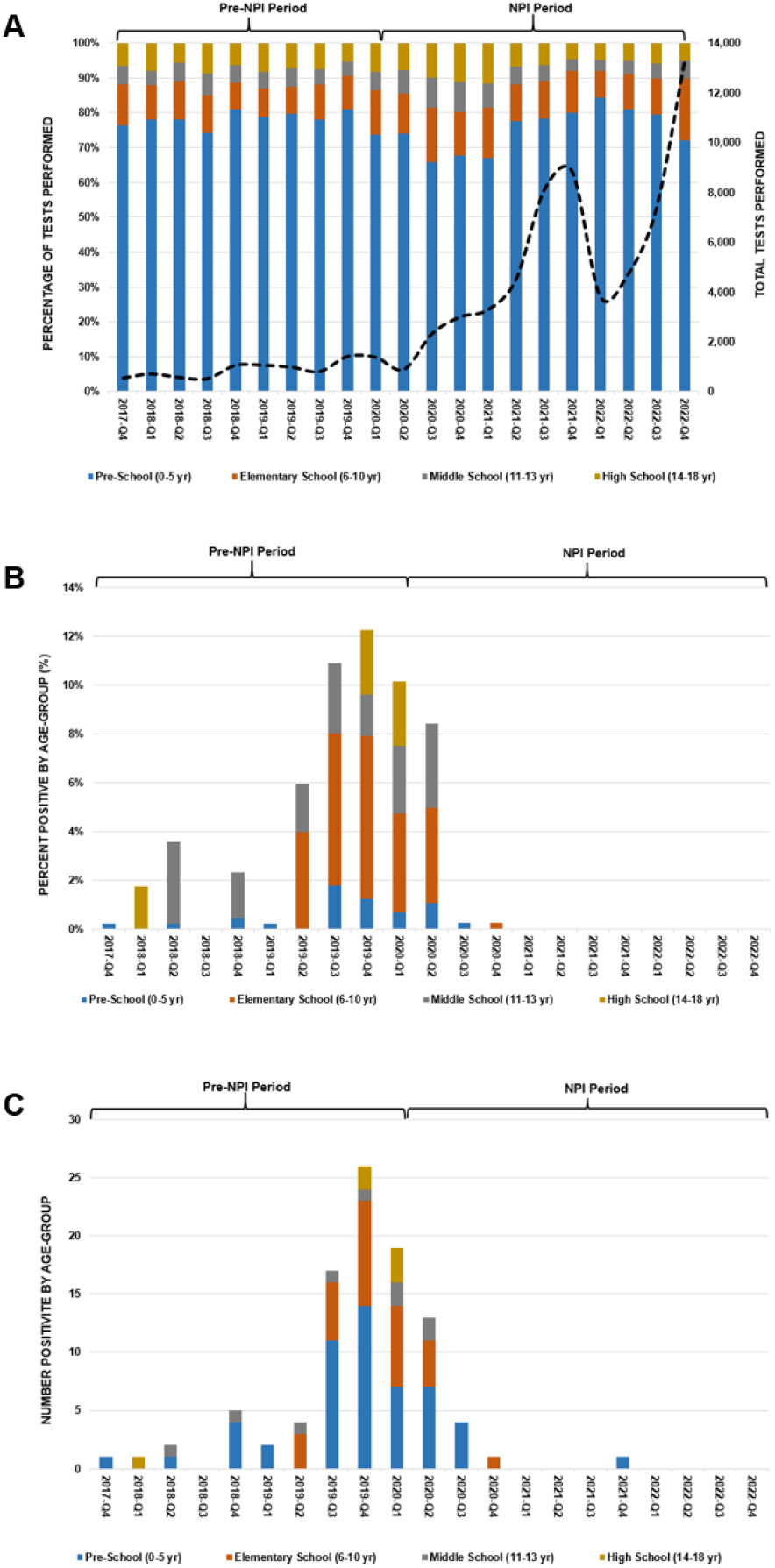
Summary of FilmArray® Respiratory Panel Testing. **(Panel A):** Total number of tests performed (black dashed line) and percentage of tests performed by age group. **(Panel B):** *M. pneumoniae* positivity rate by age group. **(Panel C):** Total number of *M. pneumoniae* positive tests by age group.

Table 2 highlights the differences in the number of coinfections (positive for *M. pneumoniae* and other upper respiratory tract bacteria and/or viruses) between the pre-NPI (n=36) and NPI (n=6) periods (p<0.001). Human rhinovirus/enterovirus was the most common coinfection among all age groups, primarily observed in the preschool and elementary school age groups. Respiratory syncytial virus, adenovirus, human metapneumovirus, parainfluenza virus, influenza virus, and non-SARS-CoV-2 coronavirus were also identified as coinfections in specific age groups. Middle school students did not exhibit any additional coinfections, while high school students had *M. pneumoniae* coinfections with adenovirus and influenza. Coinfections with two or more respiratory viruses were primarily observed in the preschool age group (n=10).

**Table 2.** Summary of Coinfections (*M. pneumoniae* and other respiratory viruses).

| School Group (Age) | Upper Respiratory Tract Coinfections |  |  |  |  |  |  |  | Pre-NPI Period (11/2017-03/2020) | NPI Period (04/2020-12/2022) |
| --- | --- | --- | --- | --- | --- | --- | --- | --- | --- | --- |
|  | Mp | RV/EV | RSV | CoV | PIV | Flu | ADV | hMPV |  |  |
| **All** |  |  |  |  |  |  |  |  | **36** | **9** |
| **Pre (0-5 yr.)** | X | X |  |  |  |  |  |  | 10 | 2 |
|  | X | X | X |  |  |  |  |  | 5 | 0 |
|  | X |  |  | X |  |  |  |  | 2 | 0 |
|  | X | X |  |  |  |  | X |  | 1 | 3 |
|  | X | X | X |  | X |  |  |  | 1 | 0 |
|  | X |  | X |  |  |  |  |  | 1 | 0 |
|  | X |  | X |  |  |  | X |  | 1 | 0 |
|  | X |  | X |  | X |  |  |  | 1 | 0 |
|  | X |  |  |  |  | X |  | X | 1 | 0 |
|  | X |  |  |  |  |  |  | X | 1 | 0 |
|  | X |  |  |  |  |  | X |  | 0 | 1 |
| **Elementary (6-10 yr.)** | X | X |  |  |  |  |  |  | 4 | 2 |
|  | X |  |  |  |  |  | X |  | 2 | 1 |
|  | X | X |  | X |  |  |  |  | 2 | 0 |
|  | X |  | X |  |  |  |  |  | 1 | 0 |
| **Middle (11-13 yr.)** | X | X |  |  |  |  |  |  | 1 | 0 |
| **High (14-18 yr.)** | X | X |  |  |  |  | X |  | 1 | 0 |
|  | X |  |  |  |  | X |  |  | 1 | 0 |

**ADV**, adenovirus; **CoV**, human coronaviruses (not SARS-CoV-2); **Flu**, influenza viruses; **RV/EV**, human rhinovirus/enterovirus; **hMPV**, human metapneumovirus; **Mp**, *Mycoplasma pneumoniae*; **PIV**, parainfluenza viruses (1,2,3,4); **RSV**, respiratory syncytial virus.

Figure 2 illustrates the total number of positive tests for *M. pneumoniae* in the 75 counties of Arkansas during the pre-NPI and NPI periods. The data presented only represent residents of Arkansas who were tested using the FARP. Out of the total 68,949 FARP tests performed, 96.5% (66,564) were from Arkansas residents, while the remaining 3.5% (2,385) were from individuals residing in neighboring states or traveling abroad for healthcare. Only two non-resident Arkansans were positive for *M. pneumoniae* during the pre-NPI (n=1) and NPI (n=1) periods. Every county in Arkansas was represented in the dataset, ranging from 3 individuals tested to 2,541 (mean 110) tests during pre-NPI period and 13 individuals tested to 9,648 (mean 777) during the NPI period.

**Figure 2.**
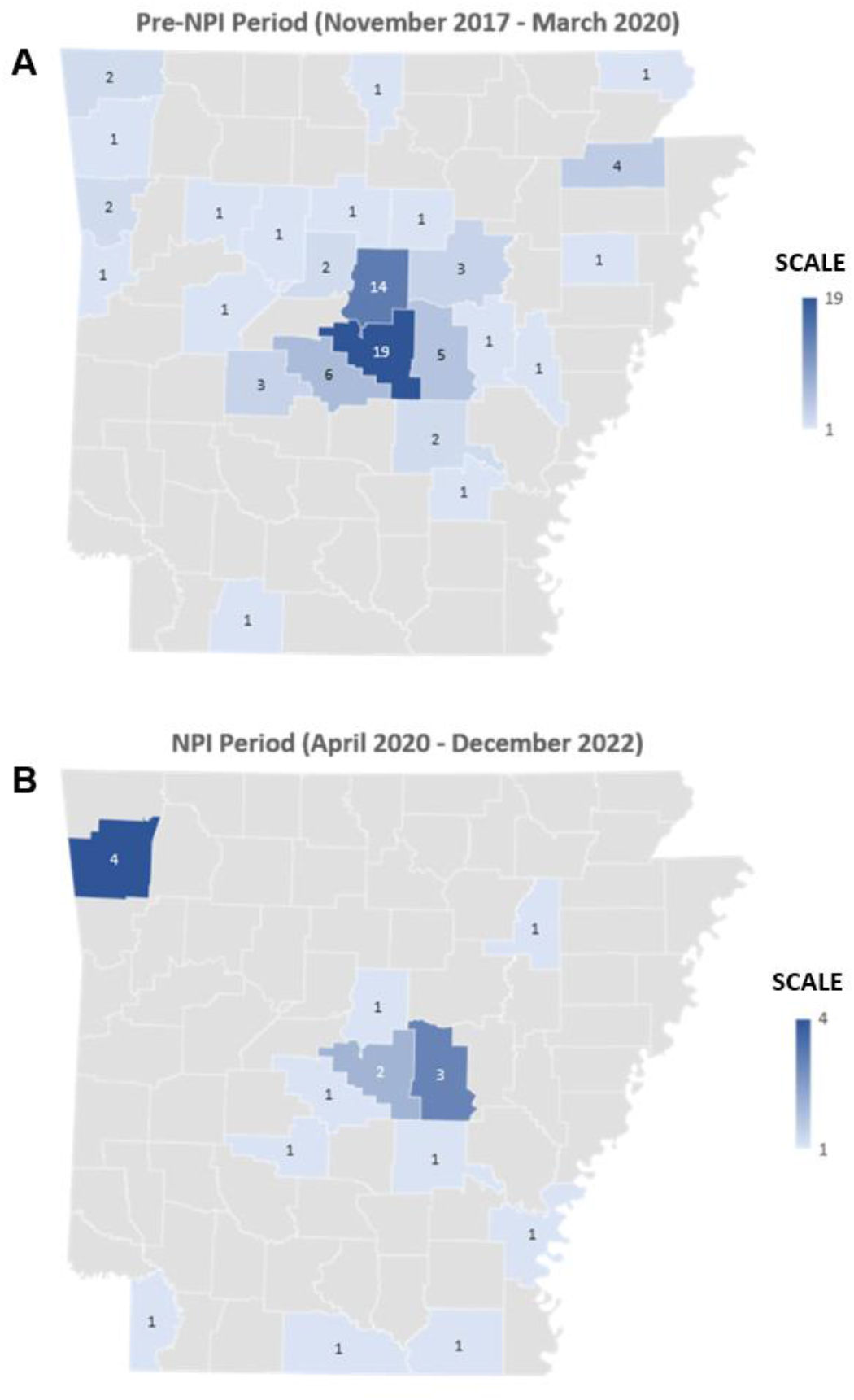
Number of Positive Tests for *M. pneumoniae* by County in Arkansas.

## Discussion

In our study, which included nearly 69,000 tests of children from every county in Arkansas, we observed a 74% reduction in *M. pneumoniae* infection rates after NPI implementation. Our data are similar to prior studies performed in Chicago, IL. (15-16) From April 2017 to March 2022, researchers demonstrated an 80% reduction in infection rates secondary to *M. pneumoniae* after implementation of NPIs (n=20,751 NAATs). Other studies across the world have shown similar decreases in *M. pneumoniae* infections. In a retrospective study from Children’s Hospital Affiliated to Capital Institute of Pediatrics (Beijing, China), 569,887 pediatric patients with respiratory infections from June 1, 2016, to May 31, 2021, were studied. (17) Using IgM-specific serology, the *M. pneumoniae* infection rate decreased from 17.59% in 2019 to 8.95% and 4.95% in 2020 and 2021, respectively. The significant decline in the *M. pneumoniae* infection rate was temporally associated with SARS-CoV-2 NPI implementation. (17)

Our study also found that the reduction in *M. pneumoniae* infections was consistent across different age groups, including preschool, elementary, middle school, and high school children. To our knowledge, these findings have not been previously reported in the United States. Similar findings were documented in Israel, China, and Finland, where *M. pneumoniae* infection rates similarly decreased during the SARS-CoV-2 pandemic.(14-21) A study from Henan Children’s Hospital (Zhengzhou, China) evaluated 1,259,697 symptomatic children (≤ 18 yrs.) from 2018 to 2021. (18) Using IgM-specific serology, the *M. pneumoniae* infection rate significantly decreased during the SARS-CoV-2 pandemic, and this was observed in all age groups (0-1 yr., 1-3 yr., 3-6 yr., and 6-18 yr.).(18) Similarly, in Finland, the number of *M. pneumoniae* infections decreased by 72-89% (NAAT) during the SARS-CoV-2 pandemic; the decrease was consistently observed in children among the 0-4 yr., 5-9 yr., and 10-14 yr. age groups (19, 21).

Data regarding children coinfected with *M. pneumoniae* and respiratory viruses predates the SARS-CoV-2 pandemic and is mostly limited to children in the United States and China with CAP. These studies, conducted between 2010 and 2019 and involving approximately 3,000 children, showed that coinfections with respiratory viruses were common, ranging from 15% to 66%. As replicated in our study, coinfections with two or more respiratory viruses were primarily observed in young children. The most frequently observed viral coinfections included human rhinovirus/enterovirus, parainfluenza viruses, adenovirus, influenza viruses, respiratory syncytial virus, human coronaviruses (excluding SARS-CoV-2), human metapneumovirus, human bocaviruses, and parechoviruses. In our study, we observed coinfections with *M. pneumoniae* and respiratory viruses in both the pre-NPI and NPI periods, with similar viral etiology and age group distribution. However, NPIs effectively reduced the transmission and acquisition of *M. pneumoniae* and most coinfecting respiratory viruses, except for human rhinovirus/enterovirus and adenovirus. It is worth noting that during the SARS-CoV-2 pandemic, NPIs successfully reduced or temporarily eliminated most circulating respiratory viruses, except for human rhinovirus/enterovirus and adenovirus, which continued to circulate at reduced levels. The reasons for this are not fully understood but may be related to factors such as asymptomatic carriage, unique transmission mechanisms, and the prolonged survival of these nonenveloped viruses on surfaces.

Our study has some limitations. First, we did not correlate positive test results with clinical, radiographic, and other laboratory information, so we cannot determine if patients were infected or colonized by *M. pneumoniae* and/or other respiratory viruses. Second, our data represents an aggregated view and may not reflect specific individuals seeking medical care in different settings. Finally, we could not verify the extent of NPI compliance among patients who underwent FARP testing during the SARS-CoV-2 pandemic, which may affect the accuracy of our data in assessing the effectiveness of NPI implementation in reducing *M. pneumoniae* and co-infecting respiratory viruses.

## Conclusions

NPIs effectively reduced the number of infections due to *M. pneumoniae* and coinfections due to *Mycoplasma pneumoniae* and most respiratory viruses in pediatric patients in the State of Arkansas. Specifically, coinfections due to *M. pneumoniae* and human rhinovirus/enterovirus and/or adenovirus were still present, albeit at lower levels, during the SARS-CoV-2 pandemic. These findings suggest that NPIs alone do not effectively eliminate these specific viruses. Due to transmission mechanisms and the nonenveloped nature of both human rhinovirus/enterovirus and adenovirus, additional infection control measures (e.g., surface decontamination and improved hand hygiene) will likely be required to reduce their spread. With the relaxation of NPIs and the recent declaration of the end of the SARS-CoV-2 pandemic, respiratory viruses will continue to return to pre-pandemic levels and establish traditional circulatory patterns. It is well known that the replication time for viruses is exponentially faster than for bacteria, including *M. pneumoniae*. Therefore, one can assume that a resurgence of infections due to *M. pneumoniae* is on the horizon and will likely occur within 1-2 years.

## Data Availability

All data produced in the present study are available upon reasonable request to the authors.

## Notes

**Conflict of Interest:** All authors declare no conflicts of interest.

### Competing Interest Statement

The authors have declared no competing interest.

### Funding Statement

Dr. Joshua Kennedy reports funding by the Center for Translational Pediatric Research (NIH/NIGMS P20GM121293) at Arkansas Childrens Hospital and the Arkansas Childrens Research Institute.

### Author Declarations

This retrospective cross-sectional study received approval from the Institutional Review Board of the University of Arkansas for Medical Sciences with waiver of consent and HIPAA authorization (No. 274080).

